# Sensitive quantification of cerebellar speech abnormalities using deep learning models

**DOI:** 10.1101/2023.04.03.23288094

**Authors:** Kyriakos Vattis, Anna C. Luddy, Jessey S. Ouillon, Nicole M. Eklund, Christopher D. Stephen, Jeremy D. Schmahmann, Adonay S. Nunes, Anoopum S. Gupta

## Abstract

**Objective:** Objective, sensitive, and meaningful disease assessments are critical to support clinical trials and clinical care. Speech changes are one of the earliest and most evident manifestations of cerebellar ataxias. The purpose of this work is to develop models that can accurately identify and quantify these abnormalities.

**Methods:** We use deep learning models such as *ResNet 18*, that take the time and frequency partial derivatives of the log-mel spectrogram representations of speech as input, to learn representations that capture the motor speech phenotype of cerebellar ataxia. We train classification models to separate patients with ataxia from healthy controls as well as regression models to estimate disease severity.

**Results:** Our model was able to accurately distinguish healthy controls from individuals with ataxia, including ataxia participants with no detectable clinical deficits in speech. Furthermore the regression models produced accurate estimates of disease severity, were able to measure subclinical signs of ataxia, and captured disease progression over time in individuals with ataxia.

**Conclusion:** Deep learning models, trained on time and frequency partial derivatives of the speech signal, can detect sub-clinical speech changes in ataxias and sensitively measure disease change over time.

**Significance:** Such models have the potential to assist with early detection of ataxia and to provide sensitive and low-burden assessment tools in support of clinical trials and neurological care.

## I. Introduction

Degenerative cerebellar ataxias (CA) are a group of disorders associated with progressive degeneration of the cerebellum. The etiology of CA is heterogeneous and includes hereditary ataxias such as ataxia-telangiectasia (AT), spinocerebellar ataxias (SCAs), and Friedreich’s ataxia (FRDA), as well as acquired ataxias such as idiopathic late-onset cerebellar ataxia (ILOCA) and multiple system atrophy (MSA). Patients with CA often experience symptoms of clumsiness, unsteady gait and slurred speech, and progression of symptoms over time can vary by ataxia type and by individual. The prevalence of dominant hereditary cerebellar ataxias is on average 2.7*/*10^5^ with SCA type 3 (SCA3) being the most common dominant ataxia while autosomal recessive ataxias have an average prevelance of 3.3*/*10^5^ with FRDA being the most frequent followed by AT [1].

Initial diagnosis of CAs and monitoring of disease over time currently relies on neurologist-performed assessments of motor and cognitive behavior. Clinical rating scales such as the Scale for the Assessment and Rating of Ataxia [2], the International Cooperative Ataxia Rating Scale [3], and the Brief Ataxia Rating Scale (BARS) [4] are structured, semi-quantitative scales that are used to track disease changes over time in natural history studies and clinical trials. Performance on these scales is currently used to determine whether a new therapy is effective or not. Unfortunately, assessments via such scales have limitations. The scales are subjective, depend on the rater’s experience in performing the assessment, and are limited by human perception. In order to achieve adequate intra- and inter-rater reliability, the scales are necessarily discrete and coarse. As they depend on a trained clinician, and are typically performed in-person, there is limited accessibility to assessments and they cannot be performed frequently in order to account for day-to-day fluctuations in symptoms. These imprecise assessments contribute to the challenge of measuring disease change, and contribute to long and large clinical trials, which increase costs and place high burden on patients. Finally, these tools are inadequate for assessing early stages of disease, when symptoms are minimally detectable. This limits the ability to perform clinical trials in early stage disease, which may be the most impactful time to intervene in order to prevent neurodegeneration.

The field of quantitative phenotyping attempts to fill this gap by using sensors and devices to quantify aspects of motor and cognitive behavior that are important in cerebellar ataxias [5]. There has been increasing interest in the use of inertial measurement units (IMUs) that record accelerometer and gyroscope data to quantify gait or limb movement [6]–[11] features, computer mouse tasks that assess arm motor control [12], and eye tracking devices to quantify eye movement abnormalities [13], [14]. Such data can be used to generate granular descriptions of disease phenotypes and severity, and can be extended to the home setting for accessible and frequent assessments.

Speech is a promising source for quantitative behavioral biomarkers in neurological conditions. Analysis of speech data has been used widely to detect and quantify severity of Parkinson’s disease, using machine learning techniques such as support vector machines and random forests models applied to acoustic speech features [15]–[17], k-nearest neighbor models [18], parallel neural networks [19] as well as gradient boosting classifiers [20]. Deep learning approaches such as bidirectional long-short term memory models [21] have also been proposed for Parkinson’s disease. Finally, in the context of amyotrophic lateral sclerosis (ALS) mixed-effects models were incorporated as discussed in [22].

Speech changes have been clinically characterized extensively in ataxias [23]–[25], and have been shown to be an early disease feature in SCAs [26]. In particular, cerebellar dysfunction results in changes in acoustic properties of speech, and the syllable alternating motion rate is slow and irregular in its temporal pattern [27]. Computational analysis of speech in ataxias has been relatively limited. The work of [28], utilized recordings of the tongue-twister phrase ‘British Constitution’ and machine learning methods such as k-nearest neighbors to assess the severity of individuals with CA with promising results. Given the acoustic and temporal signatures of ataxic speech, we hypothesized that deep learning models trained on the time and frequency gradients of the log-mel spectrogram of speech data would learn useful representations for detecting ataxia and estimating its severity, despite the modest size of the dataset.

## II. Methods

### A. Data Collection and Participants

Data were collected from 228 total participants across 463 sessions. 203 of the sessions were performed at home, with data collected from the built-in microphone of a laptop provided to the participant or from the participant’s home computer or mobile device. For the 260 sessions conducted inperson, data were collected from a lapel-attached microphone connected to an iPad. 158/228 participants had a diagnosis of ataxia and 70/228 participants were healthy individuals acting as controls. Ataxia diagnoses included spinocerebellar ataxia (5 SCA-1, 3 SCA-2, 21 SCA-3, 7 SCA-6, 6 other SCAs), ataxia-telangiectasia (46), multiple system atrophy (8), episodic ataxia (2), autosomal recessive cerebellar ataxia (3), Friedreich’s Ataxia (3) and other types (52). The participants performed a variety of speech tasks, including describing objects and scenes as well as repeating specific syllables, with each task recorded at a sampling frequency of 44.1kHz. For this analysis we use data only from the tasks of repetitive syllables such as “la-la-la”, “go-go-go”, “me-me-me” and “pa-pa-pa”. The selection of these tasks was made to isolate the motor characteristics of speech and reduce variability due to word selection. Table I summarizes the demographic information of our sample. All participants provided written assent and/or consent, and the study was approved by the Institutional Review Board at Massachusetts General Hospital with protocol numbers of 2016P001048 (7/11/2016), 2019P002752 (1/2/2020) and 2019P003458 (4/2/2020).

**TABLE I:** Demographic information of participants. M stands for male and F for female.

|  | n | M | F | Mean Age<br>[Range] | Mean<br>BARS |
| --- | --- | --- | --- | --- | --- |
| Total | 228 | 126 | 102 | 41.5 [3 - 86] | n/a |
| Control | 70 | 34 | 36 | 29.9 [3-86] | n/a |
| Ataxia | 158 | 92 | 66 | 47.0 [5-82] | 11.6 |

### B. Pre-processing of Speech Data

The audio recordings from each participant were initially down-sampled to a sampling rate of 8kHz. A noise reduction algorithm was applied based on [29], [30], which utilizes spectral gating, a process in which an estimate of noise threshold for each frequency band is made and then used to create a mask to filter the noise from the signal. In the next step, using the Librosa library [31] on the filtered signal, we computed the Mel-frequency spectrogram which represents the spectrum of signal frequencies as a function of time. This is achieved by first decomposing the signal using a short-time Fourier transform applied to windows of size 128ms every 20ms. The resulting spectrogram was then passed through a mel-spaced frequency transformation integrated into 128 frequency bins, and the magnitude of each bin was log-transformed as shown on Fig. 1. Furthermore, the spectrogram was split into “frames” of size 1s with no overlap which produced a 51 × 128 size, with the last frame zero-padded in the time dimension. Then using the WebRTC Voice Activity Detector we assign a score in the range of 0-1 for each frame defined as the fraction of instances (bins) that voice was detected over the total number of instances. Frames with a score of less than 0.6 were discarded and the rest served as the input for the models considered in this work. Finally, all frames were globally re-scaled to values between 0 and 1 and the numerical partial derivative along the time and frequency dimensions were calculated.

**Fig. 1:**
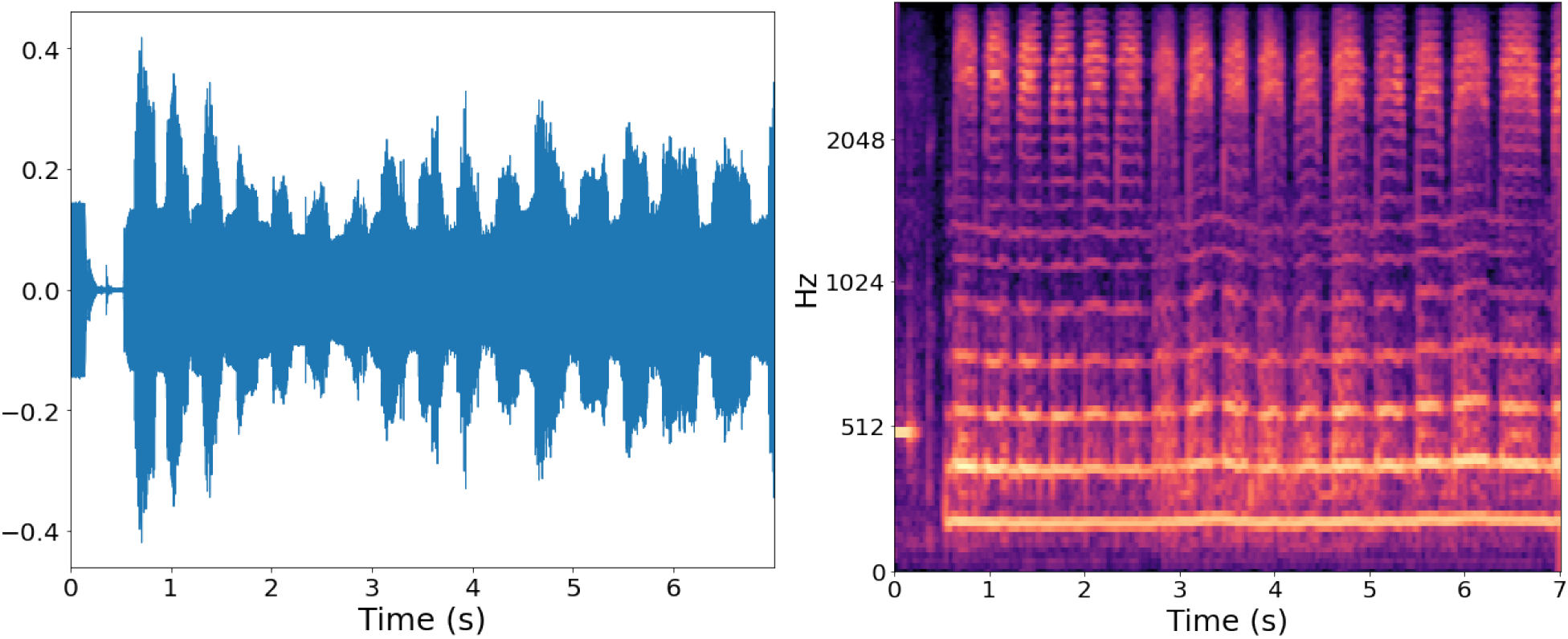
Audio sample from the repetitive task of “me-me-me” syllables. The left panel shows the waveform as a function of time while the right panel shows the resulting log-mel spectogram.

### C. Ataxia Classification

One of the objectives of this work is to create a model that uses the log-mel spectogram and its derivatives as an input to distinguish between participants with ataxia and controls. The repetitive tasks we utilize are expected to create distinct repetitive patterns in the frequency-time space as seen on Fig. 1. These patterns can occur in multiple places within the same sample and if they are general characteristics of CA, they should be present across samples from different individuals. Thus we choose to use the translational invariance of a convolutional network to take advantage of the repetitive signal.

We trained two different types of models. The first model we use is *ResNet 18* [32], a Deep Residual Network that utilizes 2-d convolutions for which we treat the spectogram inputs as monochrome images. The 2-d convolutions in *ResNet 18* are applied in the frequency-time space and can learn features that combine information across the two dimensions. The second model is a simple convolutional network with three 2-d convolutional layers that does not use the residuals like *ResNet 18*, but rather the output of the prior layer itself. Both models were trained using the Cross Entropy Loss optimized by the *Adam* optimizer [33] and their output is a probability score that the input originated from a participant with ataxia P(Ataxia). The models with parameters that minimize the loss across one epoch on the validation dataset were chosen for performance estimations.

### D. Severity Estimation

A second objective is finding a model that utilizes the speech input to estimate the ataxia severity score as measured on the BARS total score (BARS_total_) ranging from 0 - 30 as well as the speech component of the score (BARS_speech_) ranging from 0 - 4, with 30 and 4 being most severe, respectively. A modified version of BARS (BARS2) [34], which incorporates half-points was used. For this purpose, we train a *ResNet 18* model as we did for the classification task with the difference that the output is a single number representing severity. We train the model using the Mean Square Error loss optimized using the *Adam* optimizer [33] as before. The model with parameters that minimize the loss across one epoch on the validation dataset was chosen for performance estimations.

The value of BARS_total_ depends on assessments of several motor domains other than speech, including eye movements (e.g., saccades and smooth pursuit), limb reaching movements, and gait. The total score was used as one label for training, because aggregate scores tend to be more robust to errors in clinician ratings and provide higher resolution. In contrast, BARS_speech_, is the most relevant score to the tasks discussed here and is expected to better reflect severity as estimated from speech. Individuals who have normal natural speech and normal rapid production of consonants are given a BARS_speech_ score of 0. If natural speech is normal but rapid consonant production has irregularities, the individual is scored a 0.5. If there is mild slurred speech but all words are intelligible, a score of 1 is given. With increasing slurred speech and reduced intelligibility, BARS_speech_ scores increase up to a maximum score of 4 (speech is absent or unintelligible).

### E. Cross Validation

To validate our results for the classification task, we split our data into a training data set containing 80% of the ataxia participants and 80% of the controls, while the rest form a validation set. The equivalent numbers for severity estimation was 90% to ensure that representatives across the whole BARS scale were present in the training dataset. We randomly make this splitting five(ten) times ensuring that all participants pass through the validation set and train a separate model for each splitting. Choosing our datasets based on the participants and not per frame, ensured that frames that originated from the same individual were included in the same data set and there was no contamination of the validation data set with training data. The downside of this approach was that the training data set size was not exactly the same for all five(ten) folds since each participant’s task duration varies. Finally, we report the results of the five(ten) models on the same plots and performance metrics are averaged over the models.

### F. Data Augmentation

During training for the classification task, a combination of data augmentation techniques were applied to the training data set. These involved under sampling the majority class of the ataxia participants to balance the data set, as linear mix-up [35], as well as cropping along the time dimension, giving the illusion of slower speech. For the regression model, only cropping was used. As a final step all inputs, including the validation samples that did not go under the aforementioned augmentations, were resized to 100 × 100 matrices to in order to accommodate the large size of *ResNet 18*. While for the training of the models each frame was used as an independent sample, for all validation purposes the frames originating from the same participant during the same session were aggregated together and their median value is considered as the single output of the model.

## III. Results

### A. Ataxia Classification

A variety of model and input type combinations were tested and the results are summarized in Table II. The combination of *ResNet 18* with the frequency and time partial derivatives of the Mel spectrogram as an input, performed the best and was used for the plots in this section. Fig. 2 presents the output probability score as a function of BARS_total_ and BARS_speech_ with controls represented with blue markers and participants with ataxia represented with red. The model demonstrates a P(Ataxia) threshold value that well separates the two groups and both models perform well as indicated by the performance metrics as discussed below.

**TABLE II:**
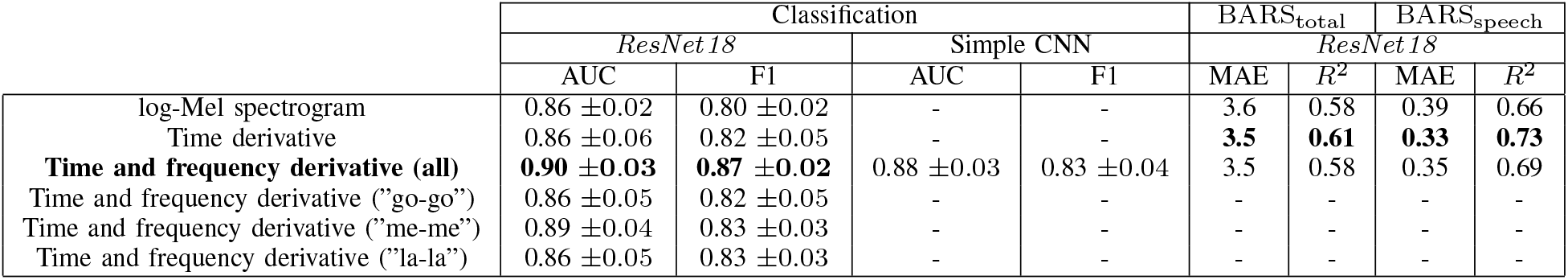
Model performance summary.

**Fig. 2:**
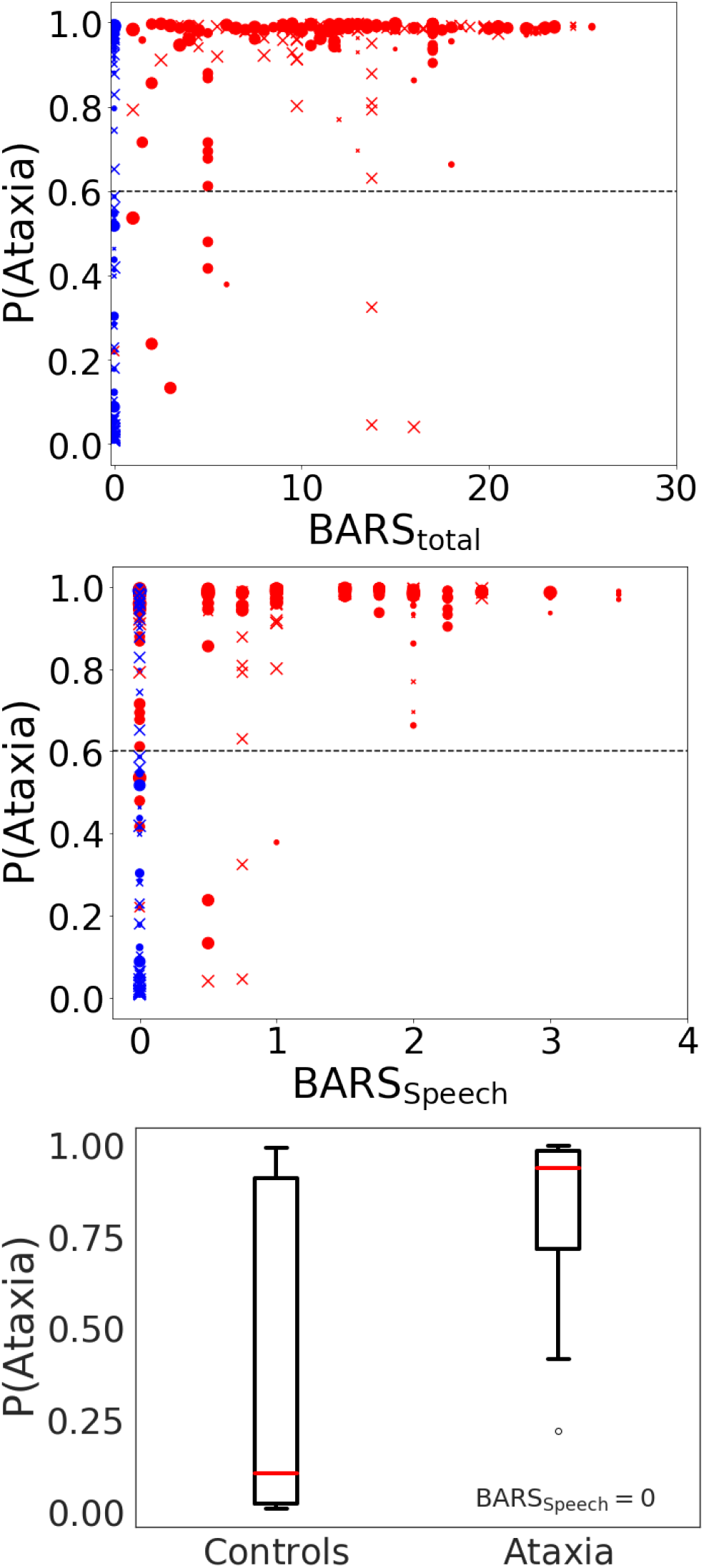
Probability of the sample originating from an individual with ataxia plotted against BARS_total_ (top) and BARS_Speech_ (middle) using the *ResNet 18* model. Blue points represent controls and red points represent participants with ataxia. The sex of the participants is identified with dots for males and crosses for females and finally the size of the plotted points increases with participants age. The dashed line is the threshold chosen for the reported metrics. At the bottom panel is the distribution of P(Ataxia) for controls and participants with ataxia and BARS_Speech_ = 0.

The mean area under the ROC curve (AUC) was 0.90±0.03 for *ResNet 18* (Fig. 3). Assuming an ataxia probability threshold for positive classification of P(Ataxia) *>* 0.6, *ResNet 18* achieved a class weighted F1 score with mean value across all 5 validation folds of 0.87 ± 0.02. The normalized confusion matrix for this threshold is shown on Fig.4.

**Fig. 3:**
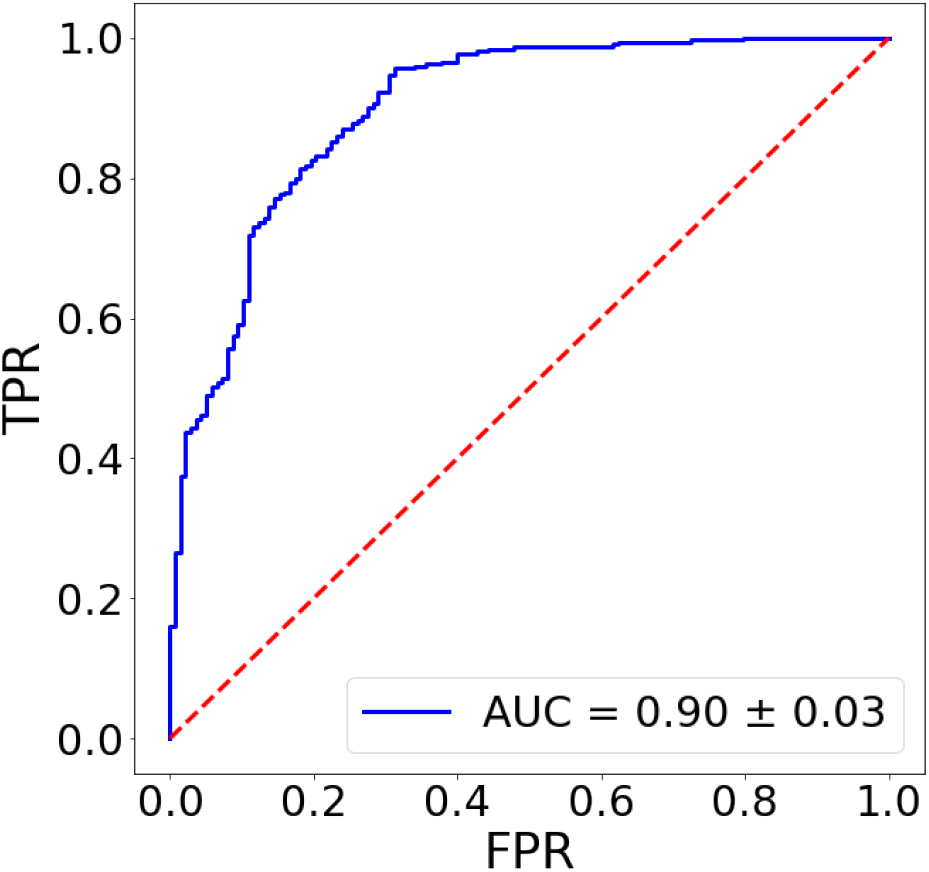
ROC curve for the classification models.

**Fig. 4:**
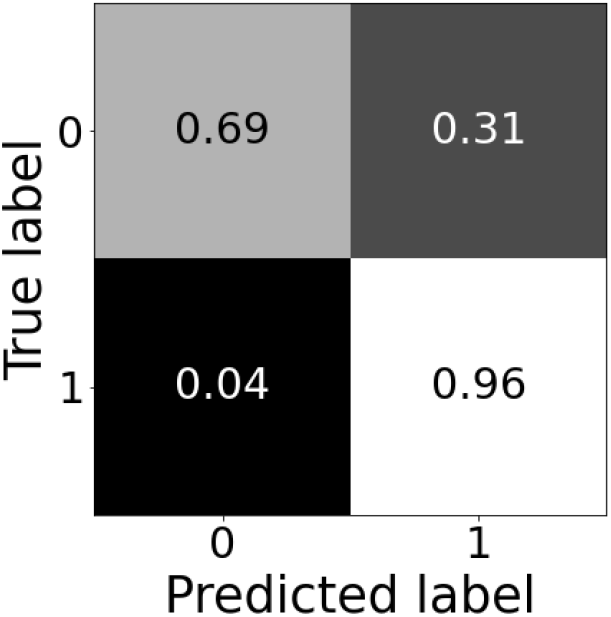
Confusion matrix for the model of the classification task with 0 labeling control and 1 labeling ataxia numbers. We assume a threshold of P(Ataxia) = 0.6.

Next, we tested whether the models could capture ataxic speech changes in individuals with ataxia but no observed clinical deficits in speech. A Mann-Whitney U-test was performed to test whether the model predicted lower *P* (*Ataxia*) values for controls compared to ataxia participants with *BARS*_*speech*_ = 0, resulting in a p-value of 6 × 10^*−*8^. This provides strong evidence that the model is capturing ataxic speech features that are not detected on the neurologist’s clinical assessment with the median *P* (*Ataxia*) = 0.10 for controls and *P* (*Ataxia*) = 0.94 for ataxia participants as seen on Fig. 2. The F1 classification score for this subset of our cohort (controls and ataxia participants with *BARS*_*speech*_ = 0) is 0.75.

Furthermore, we tested performance of the models with respect to participant sex and age. A Mann-Whitney U-test was used to compare model performance for male versus female, separately for the control and ataxia populations. We found that the model predicted higher *P* (*Ataxia*) values for males than females in the control (p = 0.02) and ataxia (p = =0.04) groups. This may in part be due to the fact that our ataxia cohort contains more males than females that could lead to some male characteristics been considered as ataxic by the model. Finally, Spearman correlation was used to evaluate the relationship between participant age and 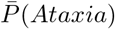 among the subjects with ataxia. The Spearman correlation coefficient was 0.09 with a p-value of 0.15, thus there was no significant relationship observed. This is further supported by the F1 and AUC metrics for various age groups as summarized in Table. III.

**TABLE III:** F1 and AUC scores for various age ranges for *ResNet 18*. N is the number of participants in that age range.

| Age [years] | F1 | AUC | N |
| --- | --- | --- | --- |
| 0 - 20 | 0.88 | 0.94 | 58 |
| 21 - 40 | 0.85 | 0.97 | 37 |
| 41 - 60 | 0.87 | 0.88 | 55 |
| > 60 | 0.94 | 0.93 | 64 |

Finally, we test the dependence of the output on ataxia severity by splitting the ataxia population into two groups, one with *BARS*_*total*_ *<* 15 and one with *BARS*_*total*_ *>*= 15. Running a Mann-Whitney U-test again, the more severe group had a higher 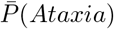 with p-value of 7 × 10^*−*3^. Splitting the dataset into a group with *BARS*_*speech*_ *<* 1.5 and one with *BARS*_*speech*_ *>*= 1.5 also demonstrated that the more severe group had a higher 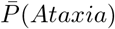 with p-value of 7 *×* 10^*−*5^.

### B. Severity Estimation

The severity estimation task was modeled as described in Methods (II-D). For this task, the combination of *ResNet 18* with time partial derivatives of the Mel spectrogram as the input (excluding frequency partials), performed the best and was used for the plots in this section. We train two separate *ResNet 18* models that predict BARS_total_ and BARS_speech_ score. The predicted BARS scores (BARS^pred^) from each of the two models are shown against the clinical BARS score (BARS^clin^) on Fig. 5. Aggregate validation data from all ten realizations of each model are plotted together. Our predictions of 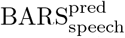 demonstrated strong agreement with the clinical scores with a Mean Absolute Error (MAE) of 0.33 and an *R*^2^ score of 0.73. Similarly, for 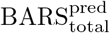, the agreement between predicted and clinical scores was MAE = 3.5 and *R*^2^ = 0.61, which is comparable with other approaches using wearable inertial measurement unit data [6].

**Fig. 5:**
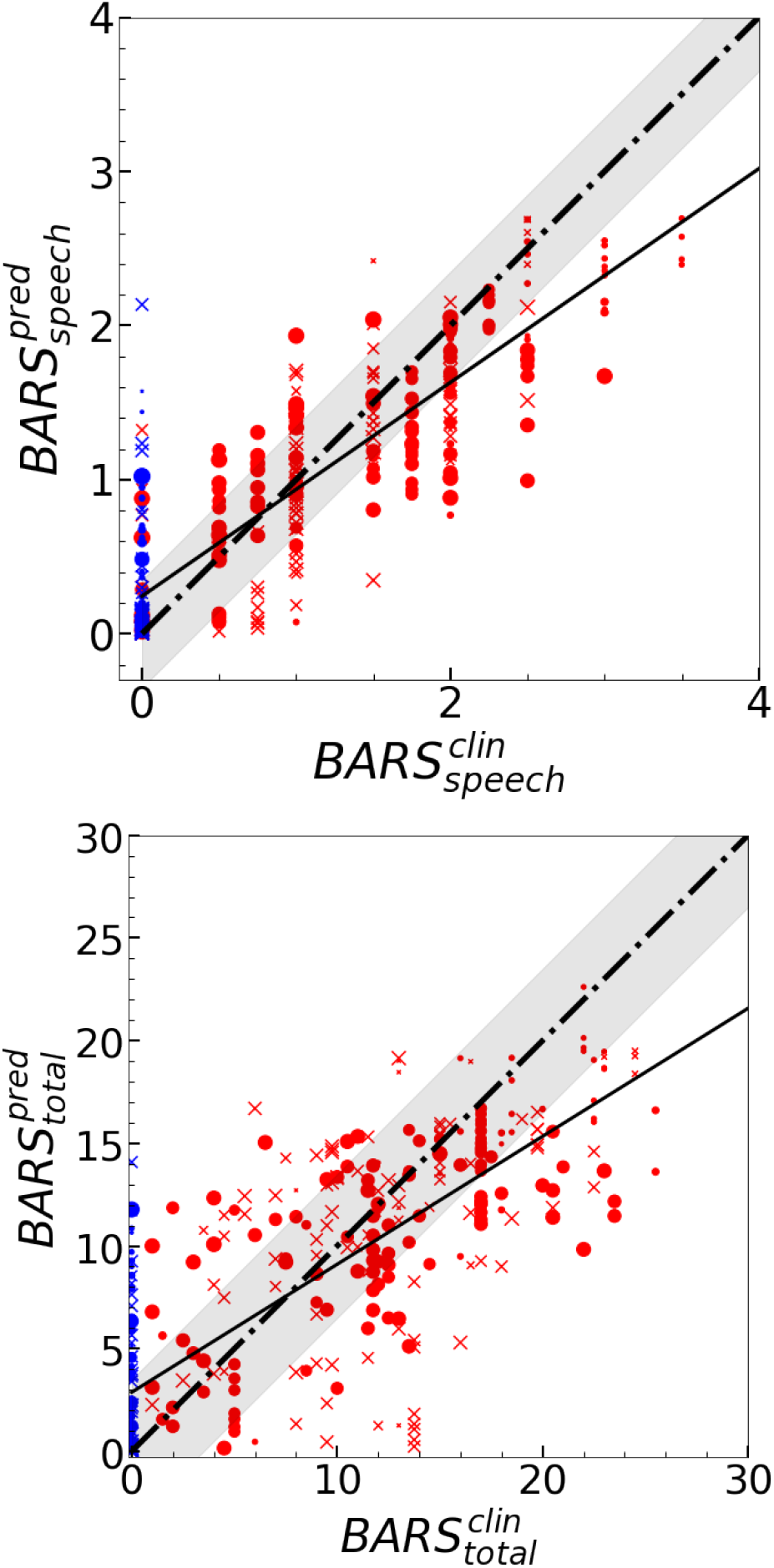
BARS score inference. The data points are modeled as on Fig. 2 and the diagonal dot-dashed line is the line of perfect agreement between predicted and clinical score, the solid line the best fit and the grey band shows the MAE interval. On the top we show the BARS total score while on the bottom we show the BARS speech score inference.

To test the sensitivity of the score estimation models to capture speech changes in individuals with very mild speech severity, we tested whether there were differences in model estimated scores for individuals with 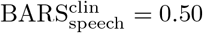 versus 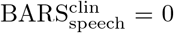. A Mann-Whitney U-test was performed to compare the population of ataxia subjects with 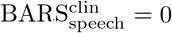 and 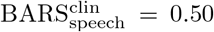 (see Fig. 6) resulting a p-value of 2 × 10^*−*4^). Thus, the models could distinguish between small variations in severity, even in very mild individuals. To further test the regression models’ ability to capture early speech characteristics, we compared 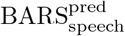 for controls and individuals with ataxia and 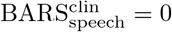. The Mann-Whitney U-test did not show a preference for lower estimates of BARS_speech_ for controls (*p* − *value* = 0.09). However, it is worth noting that the *ResNet 18* model with both time and frequency derivatives as the input did show significant model predictions between the two groups. For that model, the result shows a preference for lower estimates of BARS_speech_ for controls (*p −value* = 0.005) with a median 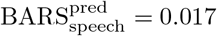 for controls and a median 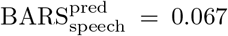 for ataxia participants. The distributions from both models are shown of Fig. 6 Finally, there is a the tendency for underestimating the scores of individuals with higher severity. This effect is especially visible in the case of 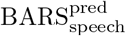 and may be attributed in part to lack of data in this high BARS_speech_ range.

**Fig. 6:**
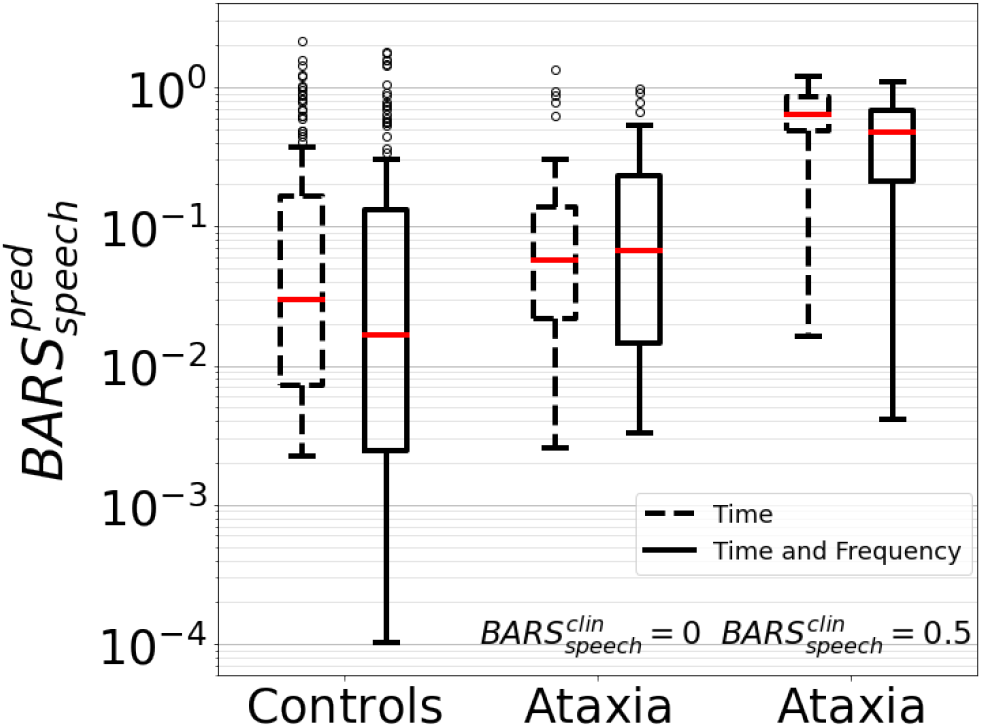
The distribution of the samples for controls on the left, ataxia participants with 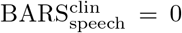 in the center and 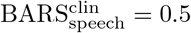 on the right. Dashed lined boxes show the distribution from the model using only time partial derivatives as input, while solid lined boxes is from the model using both time and frequency partial derivatives.

Next, we tested if the regression models could capture severity progression over time. On Fig.7 we show the change in the severity score prediction for individuals with repeated sessions. Individuals with ataxia demonstrated a statistically significant increase in predicted scores at time point 2 (p-value = 8 × 10^*−*3^ for BARS_speech_, p-value = 2 × 10^*−*2^ for BARS_total_, one-sample t-test). This demonstrates that the regression models detected progression in speech severity at subsequent visits. The Spearman correlation between change in predicated score and time between repeated assessments was 0.45 and a p-value of 9 × 10^*−*3^ for BARS_total_ and 0.46 and a p-value of 8 × 10^*−*3^ for BARS_speech_. This indicates that the models captured larger speech differences over larger time intervals, as would be expected in neurodegenerative diseases.

**Fig. 7:**
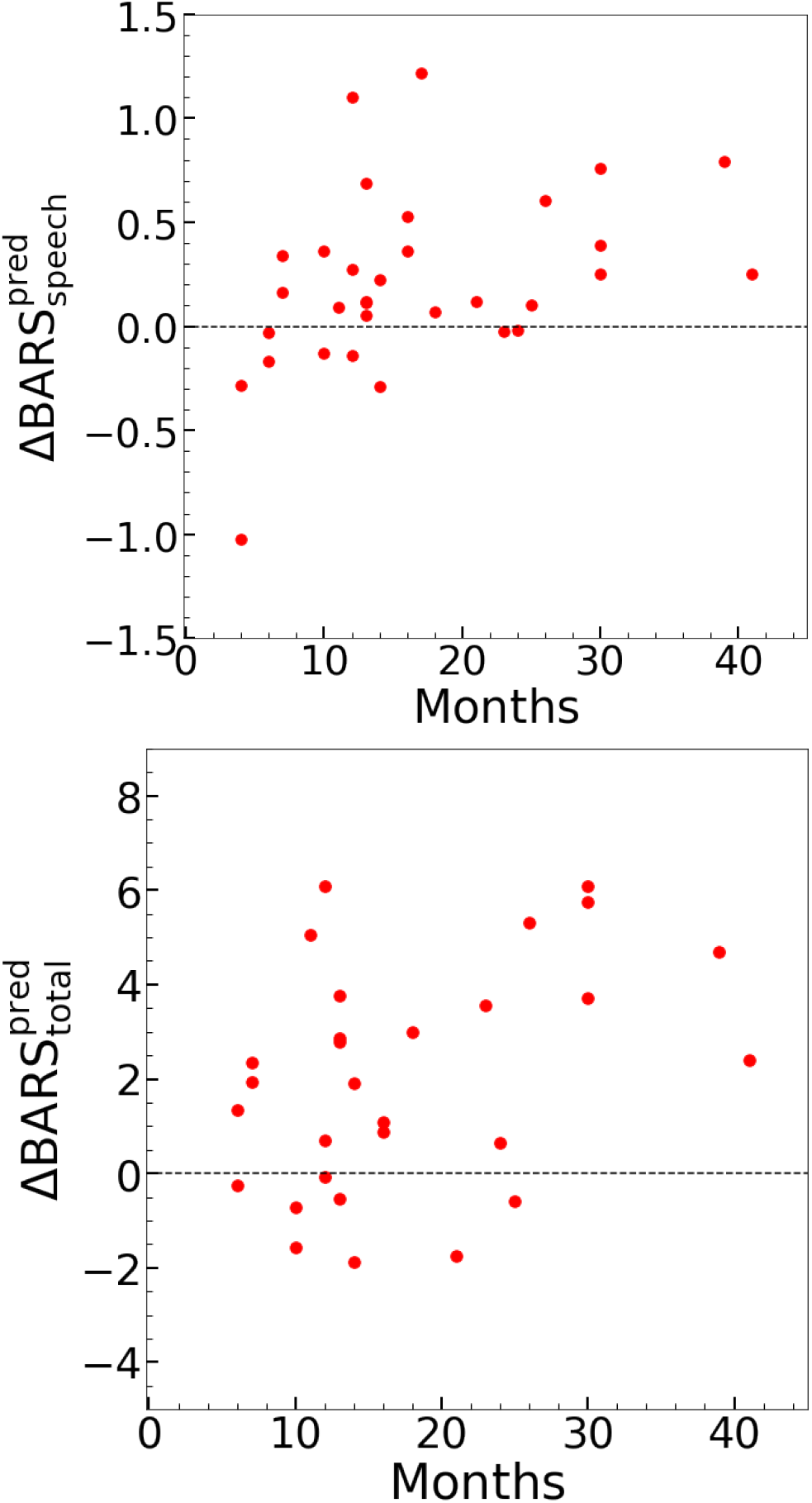
Change in the output BARS_speech_ and BARS_total_ for participants with multiple sessions spaced at least a month apart. The changes were calculated using the first and last session in record.

There was no observed dependence between participant sex and model performance for predicting *BARS*_*speech*_ in our ataxia cohort. A Mann-Whitney U-test on the distribution of Absolute Error (AE) of the *BARS*_*speech*_ predictions between male and female returned a p-value of 0.42. For *BARS*_*total*_ predictions the p-value was 0.02 for the ataxia population indicating a possible dependence on sex.The male population in our cohort had an average clinical BARS total score of 9.7 while the female population had an average of 8.0 which, along with more male participants in the dataset, may contribute to the smaller AE in the male population. No correlation was observed between AE and age for *BARS*_*speech*_ with a Spearman correlation coefficient of 0.1 and a p-value of 0.07. In contrast we observed a small correlation between AE and age for *BARS*_*total*_ with a coefficient of 0.21 and p-value of 3 × 10^*−*5^. Based on these results, the severity estimation models performed similarly well across age groups.

### C. Model Interpretability

To obtain a better understanding for the speech information utilized by the model to make predictions, Integrated Gradients (IG) [36] were calculated for correctly classified samples during test time for the classification model. We obtained one set of IG for each input component, one for the time partial derivatives and one for the frequency derivatives, which we study separately. After taking the absolute value, the Integrated Gradients of each type of derviatives, were aggregated once along the frequency dimension and once along the time dimension to obtain a salience score for each bin in time and frequency respectively. In order to visualize the IG with respect to the speech sample, Principal Component Analysis (PCA) was performed on each mel spectrogram sample. The first principal component, capturing the oscillatory behavior of the repeated syllable speech task, was used to set the boundaries for single syllables. An example of the time partial derivative-based IGs and the frequency partial derivative-based IGs for a single speech sample are shown on Fig. 8. The frequency aggregated IGs for each type of IG are overlaid in red along with the first principal component in white in order to visualize the part of the speech input in time that is contributing to correct model predictions.

**Fig. 8:**
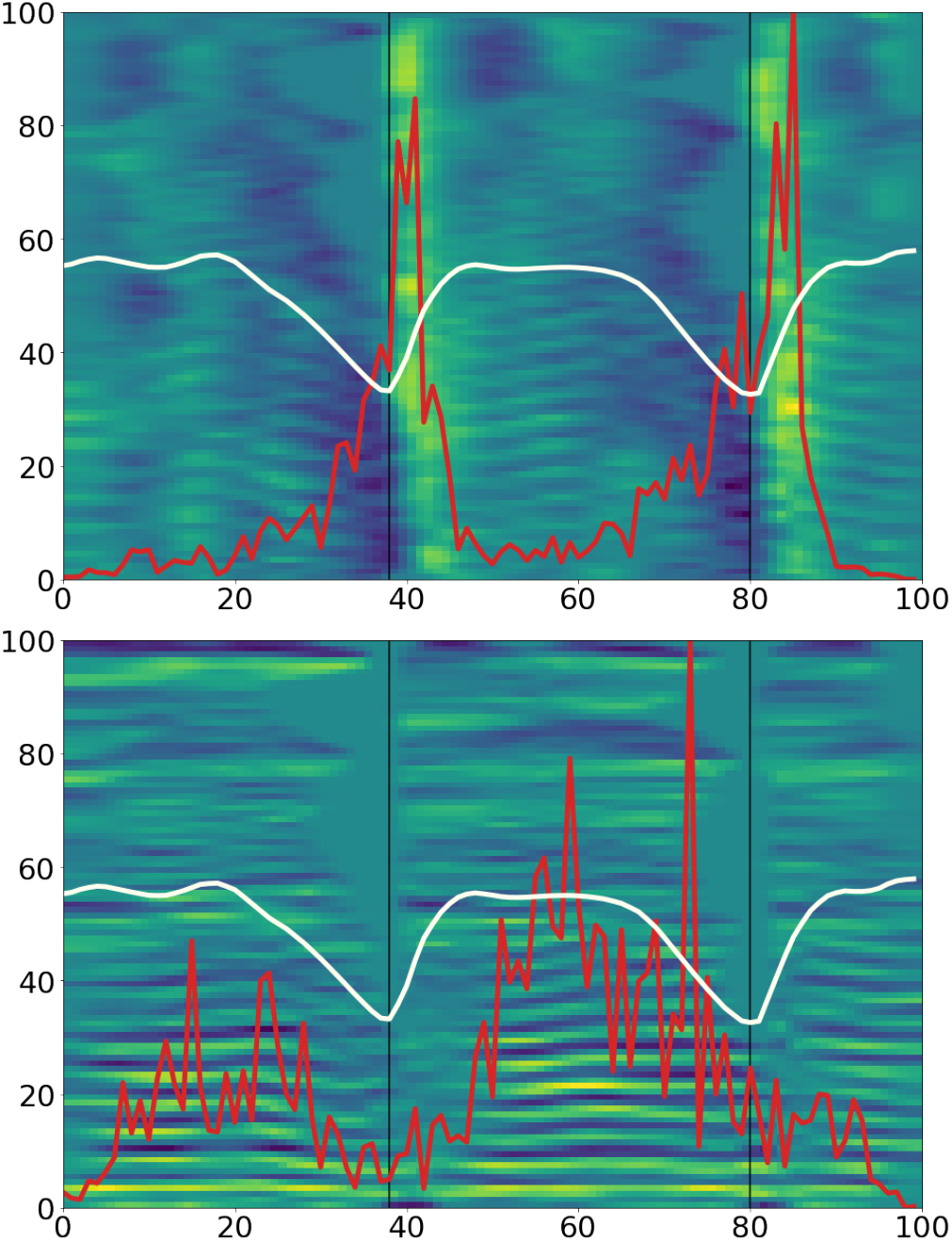
On the top panel we show a sample of mel-spectrogram time gradients where positive values are shown in green and negative values in blue. The first principal component of the spectogram is overlaid using an ivory line, the frequency aggregated salience score with red and the vertical black lines indicate the beginning and end of a syllable. We show the same for the frequency gradients on the bottom panel.

Next, an analysis was performed to determine if certain components of syllables (in time and in frequency) were preferentially informative in generating model predictions. Starting with the frequency-aggregated IG, we identify the location in time of the three instances with the three highest salience scores and measure their position in time with respect to the syllable start and end, normalized to a range of 0-1. We follow a similar procedure with the time-aggregated IG, although in this case the location of the three highest peaks is identified in frequency space, with range of 1-100.

The distribution of these peak locations across all samples is shown on Fig. 9, which reveals a structured pattern. Firstly, as shown on the the top left panel, the time variations captured by the time partial derivatives are most important at the beginning of the syllable and to a lesser extent at the end. This may be expected, as these parts of the syllables with rapid temporal change may be especially informative in ataxia, where there is known to be slow syllable alternating motion rate and irregularities in the rhythmicity. If we now turn our attention to the bottom left panel of Fig. 9 and the temporal importance of the frequency partial derivatives, we see that the distribution has a peak in the middle of the syllable. This indicates that mel spectrogram frequency gradients are most important for classification in the middle of the syllable. This could be explained as the model is using frequency-based acoustic properties of the syllable at it’s midpoint in order to make accurate predictions. Finally, the plots on the right column show that low frequencies are predominantly the most important part of the spectrum, although the time partial derivatives distribution over frequencies has a larger tail over higher frequencies and potentially a second peak (Fig. 9, top right panel).

**Fig. 9:**
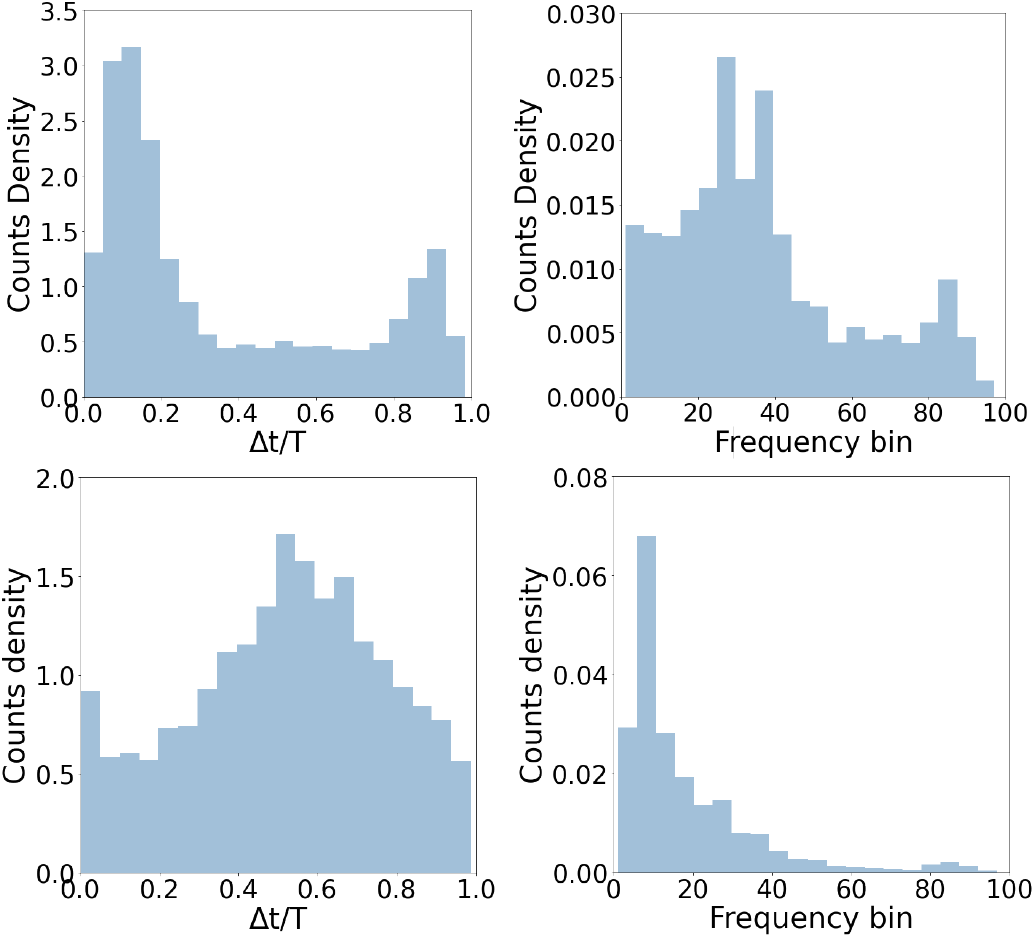
Left column: The distribution of the location of instances with maximum frequency-aggregated salience score with respect to their closest minimum of the first principal component on the left. Right column: The distribution of the location of instances with maximum time-aggregated salience score per frequency bin. Top row is using the time gradients while the bottom one uses the frequency gradients.

## IV. Discussion

Speech changes can be a powerful signal for diagnosis, severity estimation and progression measurement in cerebellar ataxias. Alternating repetitive tasks in speech and other motor domains have been widely used by neurologists to aid their assessment of ataxia patients. Thus, we leveraged the translational invariance of convolutional neural networks to capture disease-relevant information from repetitive speech task data. We have shown that a *ResNet 18* model, trained using the time and frequency partial derivatives of the log-mel spectograms of speech recordings, can accurately classify individuals with CAs and healthy individuals, including individuals without clinical speech impairment. Furthermore, we found that similarly trained *ResNet 18* models could accurately and sensitively estimate speech and total clinical severity and capture disease progression over time.

The classification models demonstrated the ability to capture sub-clinical information by accurately distinguishing ataxia participants without clinical speech deficits from controls. Such a property raises the possibility of including this type of speech analysis in a screening tool to achieve earlier diagnosis and intervention. The regression models also demonstrated sensitivity to capture subtle speech differences in very mild individuals with a clinical speech score 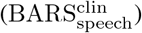 of 0 versus 0.50. A score of 0.5 indicates that natural speech is normal but that there are slight irregularities during the rapid consonant production task, whereas a score of 0 indicates that there are no detectable abnormalities in speech. The same model also produced a higher score for individuals with ataxia and 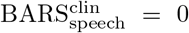 compared to controls. These results support that our approach for identifying and measuring speech changes has the potential to support early detection of ataxias, as well as the potential to produce sensitive speech measures for clinical trials.

One of the limitations of using deep learning models to make predictions, especially in health care settings, is a lack of understanding of the information leveraged by the model. As a step toward addressing this limitation for our models, we utilized integrated gradients separately for the time partial derivatives and frequency partial derivatives parts of the input. For the classification task, we found that time partial derivatives are most important at the beginning and at the end of the syllable, while frequency partial derivatives are most important in the middle. This matches intuition that the most temporally dynamic parts of the syllable may carry information about speech frequency changes in time, and the less dynamic parts encapsulate frequency-based acoustic properties of the syllables, all of which are informative in classifying individuals with ataxia.

Another key feature of this technique is its scalability. Our study included data from multiple contexts, with data collected in-person or at home, with a computer or mobile device, and using a built-in microphone or lavalier. Thus, this assessment technique that only requires participation in a repetitive speech task for less than a minute may be performed at home using everyday technologies. The ability to obtain assessments at home makes more frequent and longitudinal data collection possible and facilitates participation independent of geographical location.

A variety of other techniques and technologies have been used and proposed in the past, including inertial measurement units (IMUs) that record accelerometer and gyroscope data to quantify gait or limb movement [6], [7], computer mouse tasks that assess arm motor control [12], and eye tracking devices to quantify eye movement abnormalities [13]. Our approach complements prior work, by using convolutional neural networks with mel spectrogram time and frequency gradient inputs to probe the effects of ataxia on vocal motor impairments in an unbiased fashion. Our model performed similarly to another recent speech analysis model that was based on complex tongue twisters instead of the repetitive syllable task used here [28]. We build upon prior work by showing that in addition to generating accurate measurements of severity, carefully crafted deep learning models can learn sub-clinical information from modestly-sized speech datasets. In the future, we will seek to combine this assessment technique with quantitative assessment tools in other behavioral domains to improve on the ability to identify early signs of disease and quantify disease changes over time.

There were some limitations to the study. Most importantly, the distribution of the population of CA and control subjects in our dataset does not reflect the population distribution. In our dataset controls were the minority group while in reality the opposite is true. While we were able to account for this fact while training the models, our validation set still suffered from this imbalance and thus our classification performance estimates may be inflated compared with real-world performance. In general, while our dataset is on the larger side for studies in the field, our models would benefit from even larger datasets for training. We were also unable to account for cultural and geographical biases, such as language and accents, although we expect the single syllable repetitive tasks to be less sensitive to those factors.

## V. Conclusion

To conclude, speech changes are one of the earliest and most evident manifestations of cerebellar ataxia. We trained convolutional neural networks, with time and frequency partial derivatives of the speech signal as input, that were able to accurately separate patients with ataxia from healthy controls, including ataxia participants with no detectable clinical deficits in speech. We also trained similar regression models that could provide accurate and sensitive estimates of speech severity. We showed that integrated gradients could be used to understand how the networks use information about different parts of the syllable to generate predictions. Such speech analysis tools have the potential to assist with early detection of ataxia, to provide low-burden monitoring tools for neurological care, and to provide speech outcome measures for use in natural history studies and interventional trials. Further work is needed to fine tune and validate these models on larger and more balanced datasets that better reflect the true distribution of the population.

## Data Availability

All data produced in the present study are available upon reasonable request to the authors

## VI. Acknowledgements

The authors thank Michael Brenner for useful conversations. This project utilized resources at the Martinos-MLSC cluster. This project was funded by NIH R01 NS117826, University of Pennsylvania Orphan Disease Center in partnership with the Ataxia-Telangiectasia Children’s Project, and Biogen Inc.

## VIII. Author Contribution

K.V., and A.S.G. contributed to the conception and design of the study; A.C.L, J.S.O, N.M.E, C.D.S., J.D.S., and A.S.G. contributed to the acquisition of the data; K.V., A.S.N., and A.S.G contributed to the analysis of the data; K.V., and A.S.G. contributed to drafting the text and figures; all authors revised the manuscript for intellectual content.

## IX. Data Availability

Data could be made available upon reasonable request.

## References

[1] L. Ruano, C. Melo, M. C. Silva, and P. Coutinho, “The global epidemiology of hereditary ataxia and spastic paraplegia: a systematic review of prevalence studies,” Neuroepidemiology, vol. 42, no. 3, pp. 174–183, 2014.

[2] T. Schmitz-Hübsch, S. T. Du Montcel, L. Baliko, J. Berciano, S. Boesch, C. Depondt, P. Giunti, C. Globas, J. Infante, J.-S. Kang et al., “Scale for the assessment and rating of ataxia: development of a new clinical scale,” Neurology, vol. 66, no. 11, pp. 1717–1720, 2006.

[3] P. Trouillas, T. Takayanagi, M. Hallett, R. Currier, S. Subramony, K. Wessel, A. Bryer, H. Diener, S. Massaquoi, C. Gomez et al., “International cooperative ataxia rating scale for pharmacological assessment of the cerebellar syndrome,” Journal of the neurological sciences, vol. 145, no. 2, pp. 205–211, 1997.

[4] J. D. Schmahmann, R. Gardner, J. MacMore, and M. G. Vangel, “Development of a brief ataxia rating scale (bars) based on a modified form of the icars,” Movement Disorders, vol. 24, no. 12, pp. 1820–1828, 2009.

[5] A. S. Gupta, “Digital phenotyping in clinical neurology,” in Seminars in neurology. Thieme Medical Publishers, Inc., 2022.

[6] K. C. Knudson and A. S. Gupta, “Assessing Cerebellar Disorders With Wearable Inertial Sensor Data Using Time-Frequency and Autoregressive Hidden Markov Model Approaches,” arXiv e-prints, p. arXiv:2108.08975, Aug. 2021.

[7] W. Ilg, J. Seemann, M. Giese, A. Traschütz, L. Schöls, D. Timmann, and M. Synofzik, “Real-life gait assessment in degenerative cerebellar ataxia,” Neurology, vol. 95, no. 9, pp. e1199–e1210, 2020. [Online]. Available: https://n.neurology.org/content/95/9/e1199

[8] A. S. Gupta, A. C. Luddy, N. C. Khan, S. Reiling, and J. K. Thornton, “Real-life wrist movement patterns capture motor impairment in individuals with ataxia-telangiectasia,” The Cerebellum, pp. 1–11, 2022.

[9] J. Lee, B. Oubre, J.-F. Daneault, C. D. Stephen, J. D. Schmahmann, A. S. Gupta, and S. I. Lee, “Analysis of gait sub-movements to estimate ataxia severity using ankle inertial data,” IEEE Transactions on Biomedical Engineering, 2022.

[10] N. C. Khan, V. Pandey, K. Z. Gajos, and A. S. Gupta, “Free-living motor activity monitoring in ataxia-telangiectasia,” The Cerebellum, pp. 1–12, 2021.

[11] B. Oubre, J.-F. Daneault, K. Whritenour, N. C. Khan, C. D. Stephen, J. D. Schmahmann, S. I. Lee, and A. S. Gupta, “Decomposition of reaching movements enables detection and measurement of ataxia,” The Cerebellum, vol. 20, no. 6, pp. 811–822, 2021.

[12] K. Z. Gajos, K. Reinecke, M. Donovan, C. D. Stephen, A. Y. Hung, J. D. Schmahmann, and A. S. Gupta, “Computer mouse use captures ataxia and parkinsonism, enabling accurate measurement and detection,” Movement Disorders, vol. 35, pp. 354–358, February 2020.

[13] L. Velázquez-Pérez, C. Seifried, M. Abele, F. Wirjatijasa, R. Rodríguez-Labrada, N. Santos-Falcón, G. Sánchez-Cruz, L. Almaguer-Mederos, R. Tejeda, N. Canales-Ochoa, M. Fetter, U. Ziemann, T. Klockgether, J. Medrano-Montero, J. Rodríguez-Díaz, J. Laffita-Mesa, and G. Auburger, “Saccade velocity is reduced in presymptomatic spinocerebellar ataxia type 2,” Clinical Neurophysiology, vol. 120, no. 3, pp. 632–635, 2009. [Online]. Available: https://www.sciencedirect.com/science/article/pii/S1388245709000078

[14] Z. Chang, Z. Chen, C. D. Stephen, J. D. Schmahmann, H.-T. Wu, G. Sapiro, and A. S. Gupta, “Accurate detection of cerebellar smooth pursuit eye movement abnormalities via mobile phone video and machine learning,” Scientific reports, vol. 10, no. 1, pp. 1–10, 2020.

[15] M. Asgari and I. Shafran, “Extracting cues from speech for predicting severity of parkinson’s disease,” in 2010 IEEE International Workshop on Machine Learning for Signal Processing. IEEE, 2010, pp. 462–467.

[16] H. Hazan, D. Hilu, L. Manevitz, L. Ramig, and S. Sapir, “Early diagnosis of parkinson’s disease via machine learning on speech data, in 2012 ieee 27th convention of electrical and electronics engineers in israel,” Google Scholar Google Scholar Cross Ref Cross Ref, 2012.

[17] A. Tsanas, M. A. Little, P. E. McSharry, J. Spielman, and L. O. Ramig, “Novel speech signal processing algorithms for high-accuracy classification of parkinson’s disease,” IEEE transactions on biomedical engineering, vol. 59, no. 5, pp. 1264–1271, 2012.

[18] H.-L. Chen, C.-C. Huang, X.-G. Yu, X. Xu, X. Sun, G. Wang, and S.-J. Wang, “An efficient diagnosis system for detection of parkinson’s disease using fuzzy k-nearest neighbor approach,” Expert systems with applications, vol. 40, no. 1, pp. 263–271, 2013.

[19] F. Aström and R. Koker, “A parallel neural network approach to prediction of parkinson’s disease,” Expert systems with applications, vol. 38, no. 10, pp. 12 470–12 474, 2011.

[20] D. Jain, A. K. Mishra, and S. K. Das, “Machine learning based automatic prediction of parkinson’s disease using speech features,” in Proceedings of International Conference on Artificial Intelligence and Applications. Springer, 2021, pp. 351–362.

[21] C. Quan, K. Ren, and Z. Luo, “A deep learning based method for parkinson’s disease detection using dynamic features of speech,” IEEE Access, vol. 9, pp. 10 239–10 252, 2021.

[22] G. M. Stegmann, S. Hahn, J. Liss, J. Shefner, S. Rutkove, K. Shelton, C. J. Duncan, and V. Berisha, “Early detection and tracking of bulbar changes in als via frequent and remote speech analysis,” NPJ digital medicine, vol. 3, no. 1, pp. 1–5, 2020.

[23] E. Schalling and L. Hartelius, “Speech in spinocerebellar ataxia,” Brain and language, vol. 127, no. 3, pp. 317–322, 2013.

[24] E. Schalling, B. Hammarberg, and L. Hartelius, “Perceptual and acoustic analysis of speech in individuals with spinocerebellar ataxia (sca),” Logopedics Phoniatrics Vocology, vol. 32, no. 1, pp. 31–46, 2007.

[25] T. Schirinzi, A. Sancesario, E. Bertini, E. Castelli, and G. Vasco, “Speech and language disorders in friedreich ataxia: highlights on phenomenology, assessment, and therapy,” The Cerebellum, vol. 19, no. 1, pp. 126–130, 2020.

[26] A. P. Vogel, M. Magee, R. Torres-Vega, J. Medrano-Montero, M. P. Cyngler, M. Kruse, S. Rojas, S. C. Cubillos, T. Canento, F. Maldonado, Y. Vazquez-Mojena, W. Ilg, R. Rodríguez-Labrada, L. Velázquez-Pérez, and M. Synofzik, “Features of speech and swallowing dysfunction in pre-ataxic spinocerebellar ataxia type 2,” Neurology, vol. 95, no. 2, pp. e194–e205, 2020. [Online]. Available: https://n.neurology.org/content/95/2/e194

[27] R. D. Kent, J. F. Kent, J. R. Duffy, J. E. Thomas, G. Weismer, and S. Stuntebeck, “Ataxic dysarthria,” Journal of Speech, Language, and Hearing Research, vol. 43, no. 5, pp. 1275–1289, 2000. [Online]. Available: https://pubs.asha.org/doi/abs/10.1044/jslhr.4305.1275

[28] B. Kashyap, P. N. Pathirana, M. Horne, L. Power, and D. Szmulewicz, “Automated tongue-twister phrase-based screening for cerebellar ataxia using vocal tract biomarkers¡sup¿*¡/sup¿,” in 2019 41st Annual International Conference of the IEEE Engineering in Medicine and Biology Society (EMBC), 2019, pp. 7173–7176.

[29] T. Sainburg, “timsainb/noisereduce: v1.0,” Jun. 2019. [Online]. Available: https://doi.org/10.5281/zenodo.3243139

[30] T. Sainburg, M. Thielk, and T. Q. Gentner, “Finding, visualizing, and quantifying latent structure across diverse animal vocal repertoires,” PLoS computational biology, vol. 16, no. 10, p. e1008228, 2020.

[31] B. McFee, C. Raffel, D. Liang, D. P. Ellis, M. McVicar, E. Battenberg, and O. Nieto, “librosa: Audio and music signal analysis in python,” in Proceedings of the 14th python in science conference, vol. 8. Citeseer, 2015, pp. 18–25.

[32] K. He, X. Zhang, S. Ren, and J. Sun, “Deep residual learning for image recognition,” CoRR, vol. abs/1512.03385, 2015. [Online]. Available: http://arxiv.org/abs/1512.03385

[33] D. P. Kingma and J. Ba, “Adam: A Method for Stochastic Optimization,” arXiv e-prints, p. arXiv:1412.6980, Dec. 2014.

[34] H. Zhou, H. Nguyen, A. Enriquez, L. Morsy, M. Curtis, T. Piser, C. Kenney, C. D. Stephen, A. S. Gupta, J. D. Schmahmann et al., “Assessment of gait and balance impairment in people with spinocerebellar ataxia using wearable sensors,” Neurological Sciences, pp. 1–11, 2021.

[35] H. Zhang, M. Cisse, Y. N. Dauphin, and D. Lopez-Paz, “mixup: Beyond empirical risk minimization,” 2018.

[36] M. Sundararajan, A. Taly, and Q. Yan, “Axiomatic Attribution for Deep Networks,” arXiv e-prints, p. arXiv:1703.01365, Mar. 2017.

